# Impact of Selection Bias on Polygenic Risk Score Estimates in Healthcare Settings

**DOI:** 10.1101/2022.07.20.22277710

**Authors:** Younga Heather Lee, Tanayott Thaweethai, Yi-han Sheu, Yen-Chen Anne Feng, Elizabeth W. Karlson, Tian Ge, Peter Kraft, Jordan W. Smoller

## Abstract

**Background:** Hospital-based biobanks have become an increasingly prominent resource for evaluating the clinical impact of disease-related polygenic risk scores (PRS). However, biobank cohorts typically rely on selection of volunteers who may differ systematically from non-participants.

**Methods:** PRS weights for schizophrenia, bipolar disorder, and depression were derived using summary statistics from the largest available genomic studies. These PRS were then calculated in a sample of 24,153 European ancestry participants in the Mass General Brigham (MGB) Biobank. To correct for selection bias, we fitted a model with inverse probability (IP) weights estimated using 1,839 sociodemographic and clinical features extracted from electronic health records (EHRs) of eligible MGB patients. Finally, we tested the utility of a modular specification of the IP weight model for selection.

**Results:** Case prevalence of bipolar disorder among participants in the top decile of bipolar disorder PRS was 10.0% (95% CI: 8.8%-11.2%) in the unweighted analysis but only 6.2% (5.0%-7.5%) when selection bias was accounted for using IP weights. Similarly, case prevalence of depression among those in the top decile of depression PRS was reduced from 33.5% (31.7%-35.4%) in the unweighted analysis to 28.9% (25.8%-31.9%) after IP weighting. Modular correction for selection bias in intermediate selection steps did not substantially impact PRS effect estimates.

**Conclusions:** Non-random selection of participants into volunteer biobanks may induce clinically relevant selection bias that could impact implementation of PRS and risk communication in clinical practice. As efforts to integrate PRS in medical practice expand, recognition and mitigation of these biases should be considered.

## INTRODUCTION

In recent years, large-scale healthcare systems have contemplated integrating polygenic risk scores (PRS) into clinical practice given their potential to stratify diagnostic and therapeutic strategies in common medical conditions (e.g., diabetes, cancer, obesity) (1–6) and, more recently, in psychiatric conditions (7). For example, the Electronic Medical Records and Genomics (eMERGE) Network is conducting trials evaluating the impact of returning genomic results (“return of results” or RoR) in both clinical and research venues (8,9). Early evidence suggests that patients are in favor of being informed of their genetic test results and receiving advice about how to interpret and act on the results (10–12). With respect to returning psychiatric PRS results, individuals living with bipolar disorder (BD) were highly accepting of polygenic risk information for BD—even more so when accompanied by comprehensive psychiatric genetic counseling (13).

With the prospect of using PRS to guide clinical decision making, optimizing the accuracy of the risk estimates they provide becomes especially important (14). In research settings, including biobank-based studies, genetic analyses are usually restricted to individuals who have volunteered to provide biospecimens for research investigations. More specifically, application of PRS in a biobank or other research cohort typically entails a sequence of sampling procedures. First, the cohort is limited to participants who provided consent, and had blood samples drawn and genotyped prior to the time of analysis. Next, this subsample is further restricted to those who have passed a genomic quality control (QC) process. However, restricting analyses without considering the complexity of selection mechanism can change or induce spurious associations between factors directly or indirectly related to selection into the PRS analysis.

Inverse probability (IP) weighting is an established method for correcting such bias in which the contribution of each sampled individual is weighted by the inverse of their probability of being sampled (15). In most volunteer-based studies, information about those who were not enrolled is typically limited, precluding in-depth exploration of selection bias that can result from non-random sampling. However, biobanks nested within healthcare systems where demographic and clinical data are available for the full healthcare system population provide a unique opportunity to evaluate factors that may influence the probability of being selected into an analytic sample. In these settings, one can use IP weighting to construct a hypothetical population in which participants are weighted such that they represent the entire population of participants and non-participants with respect to the predictors of selection and conduct analyses that account for non-random sampling.

A key assumption of IP weighting, however, is that one has correctly identified and weighted the predictors of sampling; violation of this assumption may lead to residual or even greater bias (16). Meeting this requirement could be particularly challenging in the case of hospital-based biobanks, since selection may be dynamic and reflect a large number of poorly understood factors—including patient comorbidity profiles and the diversity of clinical settings in which recruitment was conducted. Instead of solely relying on expert knowledge to specify the weight model, Haneuse and Daniels suggest combining clinical knowledge with data-driven strategies for covariate selection (17–19), especially when working with high-dimensional electronic health records (EHRs) (20). Accordingly, we use a two-step approach to correct for non-random sampling in PRS analyses. First, we apply a machine learning approach to examine the relative contribution of sociodemographic, healthcare utilization, and clinical characteristics (captured in the longitudinal EHRs) and estimate IP weights for selection. Next, we estimate the association between PRS and the target conditions in an IP-weighted sample in which selection into the Biobank study occurred at random. Using this two-step approach, we find that standard PRS analyses that do not account for the non-random sampling of biobank samples may lead to biased estimation of polygenic risk in the context of psychiatric conditions.

Finally, we address the fact that selection into biobank-based studies typically involves multiple steps—such as recruitment, consent, biospecimen collection, genotyping, and genomic QC—each of which may be influenced by a unique set of determinants (see e**Figure 1**).

**Figure 1.**
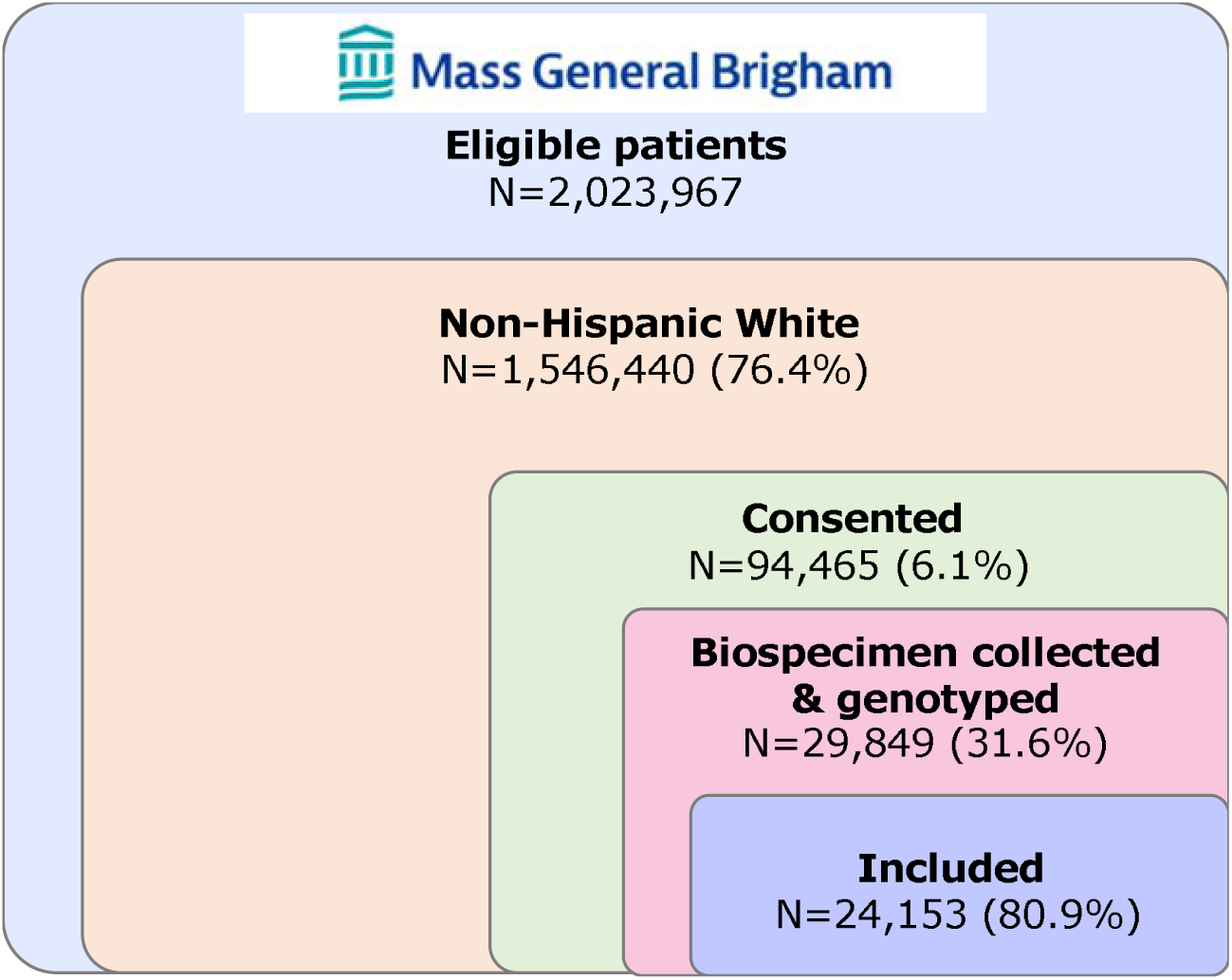
Schematic of sample curation for polygenic risk score analysis using the MGB Biobank sample.

Haneuse and Daniels proposed a general statistical framework that explicitly models the decisions made by patients and healthcare providers that collectively determine which data are available in the EHR relevant to a given research inquiry (20). They conceptualize selection bias as a missing data problem and encourage researchers to *modularize* the complex selection mechanism into a series of sub-mechanisms that are easier to characterize and model (21). Applying this modular IP weighting framework, we evaluate the discrepancy between PRS effect estimates for psychiatric conditions when using standard versus modular approaches to defining selection mechanisms.

## METHODS

### Study sample

#### Mass General Brigham (MGB) Research Patient Data Registry (RPDR)

The primary data source was the MGB RPDR, an EHR data warehouse covering 4.6 million patients across the MGB HealthCare hospital system (formerly Partners HealthCare) including Brigham & Women’s Hospital, Massachusetts General Hospital, and other affiliated hospitals in the greater Boston area. To assemble the cohort for this study, we queried the MGB RPDR for 1,546,440 patients who self-identified as non-Hispanic White (that is, 74% of the overall MGB patient population) having at least three visits after 2005, more than 30 days apart between the first and last visits, and at least one visit greater than age 10 and less than age 90, as of February 2020 (22,23) (see **Figure 1**). The race and ethnicity restriction was applied here because the subsequent PRS were based on samples of European ancestry.

#### MGB Biobank

The MGB Biobank is a hospital-based research program launched in 2010 to empower genomic and translational research for human health (12). Participants are patients at MGB-affiliated hospital(s) above age 18 (at the time of the recruitment) who provided informed consent to join the Biobank study. Each consented participant was asked to provide blood samples (e.g., plasma, serum, DNA), which are then linked to their clinical data in the EHRs as well as survey data on lifestyle, behavioral and environmental factors, and family history. Leveraging in-person and electronic recruitment methods, the MGB Biobank has currently enrolled more than 130,000 participants, collected 82,092 DNA samples, and generated genotyping microarray data for more than 56,923 participants (4,920 using the Illumina MEGA, 5,334 using the Illumina MEGA EX, 26,144 using the Illumina MEG, and 24,789 using the Illumina GSA) (23). This research was conducted as part of the PsycheMERGE Consortium (24), under approval from the MGB Institutional Review Board.

### Data-driven approach to specify IP weight models for selection

We employed a machine learning modeling method—extreme gradient boosting (XGBoost) classification (25)—to empirically identify key determinants of non-random sampling of biobank participants and calculate the IP selection weights using a large set of demographic and clinical features extracted from high-dimensional EHRs including 15 sociodemographic, 10 healthcare utilization, and 1,814 diagnostic characteristics (see **eTable 4** for the full list of features used to train XGBoost models). XGBoost is an open-source library providing a computationally efficient and high-performance implementation of gradient boosted decision trees (https://github.com/dmlc/xgboost).

In the first set of IP weighted analyses (i.e., standard IP weighted approach), we fitted an XGBoost model classifying the inclusion into the PRS analysis (n=24,153) from a pool of 1,546,440 adult patients at MGB-affiliated hospital(s) self-identifying as non-Hispanic White (7). Considering that a very small proportion of the patient population participated in the Biobank study, we ensured that the training and test sample (with a split ratio of 80:20) had the same proportion of the target outcome in a given selection step (e.g., included vs. not included in the PRS analysis for the standard IP weighting approach). After fitting the model, we derived weights by taking the inverse of the predicted probabilities of being selected into the final PRS analysis. We further stabilized the IP weights by dividing the predicted probabilities by the marginal probability of selection and truncated the top and bottom 1% of the distribution to account for extreme weights (8).

In the second set of IP-weighted analyses (i.e., modular IP weighted approach), we fit three separate sets of XGBoost models classifying each of the three selection steps (see **eFigure 1**). The three targets for classification were: 1) consent status among eligible participants, 2) biospecimen collection and genotyping status among consented participants, and 3) inclusion in the PRS dataset among participants who are eligible, consented, and had biospecimens collected and genotyped. We extracted predicted probabilities from each of the three models and took the product of these conditional probabilities to calculate the joint probabilities of being included in the final analytic sample given the three sequential steps of selection. We then stabilized and truncated the inverse of the joint probabilities in the same way as we did for the standard IP weighting approach and performed weighted PRS analyses.

In addition, we applied a game theory-based algorithm called Shapley Additive Explanations (SHAP) method (https://github.com/slundberg/shap) to further elucidate the complex selection mechanism of the MGB Biobank. We calculated Shapley values, which are the weighted average of the marginal contribution of each feature value toward the model’s decision, to explain how changes in a feature value would shift the models’ decision both in terms of absolute magnitude and direction (26). This way, we characterized the importance of each feature to the predicted probability of being retained in the study sample at each step of selection (see rankings and directionality of contribution by the top 20 features in **eFigures 2 and 3**).

### PRS construction

We generated PRS for the 24,153 MGB Biobank participants of European ancestry using their genotype data and weights derived by applying PRS-CS-Auto (27), a Bayesian polygenic prediction method, to publicly available summary statistics from the largest genome-wide association studies (GWAS) of schizophrenia (28), bipolar disorder (29), and depression (30) on populations of European ancestry (see **eMethods** for details on genomic data processing and **eTable 2** for further information on discovery GWAS).

### Case definition

We identified cases of the three psychiatric traits by mapping the entire longitudinal health records available on all patients at MGB-affiliated hospital(s) to the phecode system using the *PheWAS* R package (31,32). We identified qualifying ICD-9CM and ICD-10CM codes for schizophrenia (phecode 295.1), bipolar disorder (phecode 296.1), and depression (phecode 296.2), and defined cases as those having at least two qualifying ICD codes for a given phecode (see the full list of qualifying diagnostic codes in **eTable 3**).

### Statistical analysis

We compared effect estimates of the associations between schizophrenia, bipolar disorder, and depression PRS and their respective target diagnoses using three approaches: unweighted, standard IP-weighted, and modular IP-weighted as described below (see **eFigure 1**). In the unweighted approach, PRS effect estimates are calculated without accounting for non-random sampling. In contrast, the latter two approaches involve a systematic evaluation and adjustment for differential probabilities of being selected into the analytic sample for the PRS analyses. The application of IP weights allows us to construct a hypothetical population in which we can estimate the effects of PRS in the absence of spurious associations induced by participation-related factors specified in the weight model (see **Figure 2**). Prior to calculation of penetrance and discrimination of the PRS, we fitted linear regression models with age, sex assigned at birth, top 20 genetic principal components, and genotyping microarray as predictors of each respective psychiatric PRS. We then extracted and standardized the residuals from each regression model and generated a categorical version of the PRS using deciles. In the current study, we primarily focus on disease risk for the top decile of the standardized residuals of PRS, a threshold commonly used to define high genetic risk in the context of clinical translation (33).

**Figure 2.**
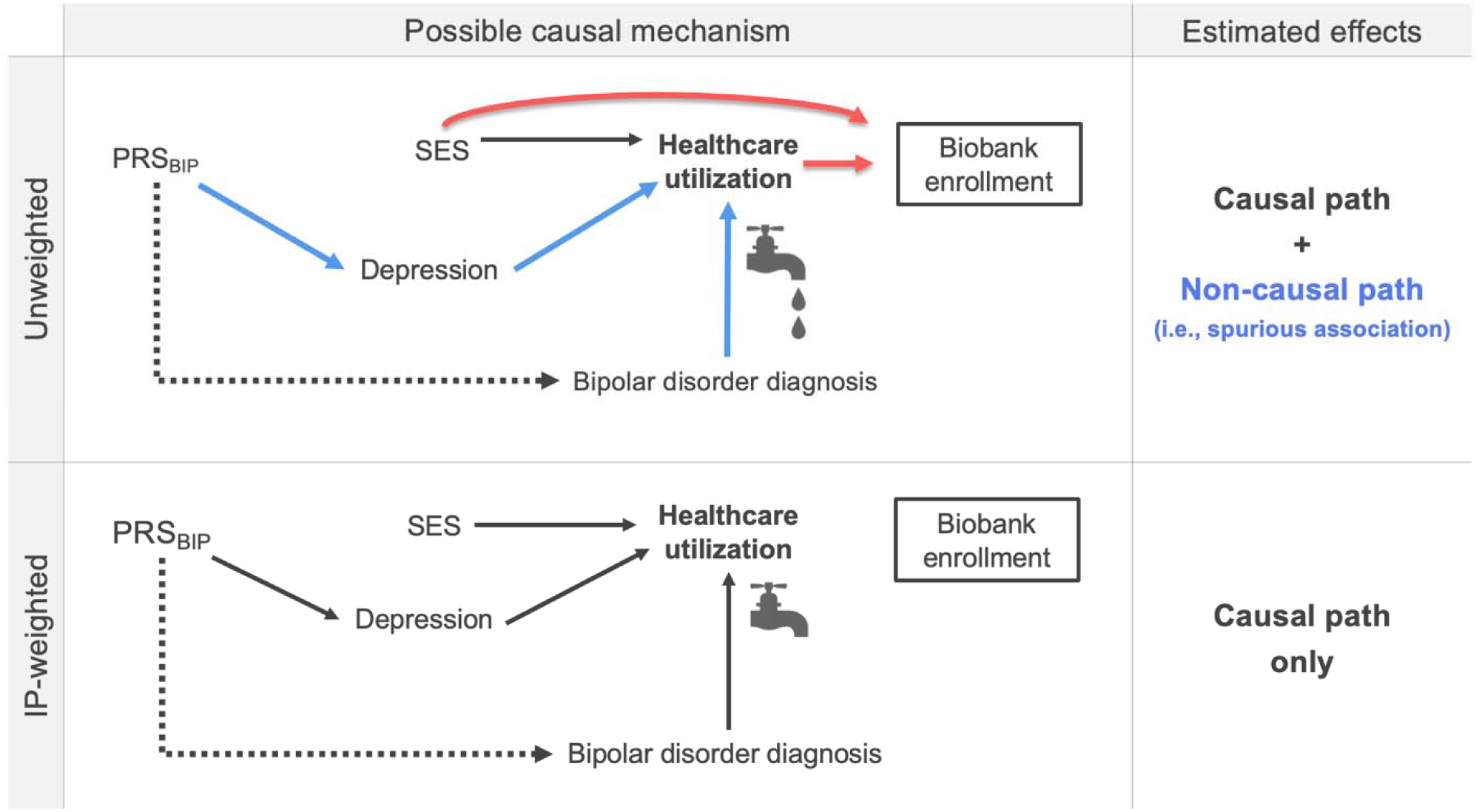
Causal diagram (directed acyclic graph or DAG) illustrating how non-random sampling into hospital-based biobanks may introduce bias in PRS estimation. Using the example of a bipolar disorder PRS, this figure depicts two DAGs to illustrate how selection bias might inflate PRS effect estimates in a hospital-based biobank in unweighted PRS analysis. The relationship of interest is denoted by the dotted line connecting PRS_BIP_ (bipolar disorder polygenic risk score) with bipolar disorder diagnosis. Restriction of PRS analysis to biobank participants is represented as a box around biobank enrollment in the causal diagram. Healthcare utilization is a common effect of PRS_BIP_ (through the effect of PRS_BIP_ on depression) and clinical diagnosis of bipolar disorder. In this example, stratification on biobank enrollment, a descendant of healthcare utilization, can induce a spurious association between the PRS and the target trait (represented as a dripping faucet in the figure below). Thus, the estimated effect could include not only true causal effects but also the spurious association, thereby resulting in larger estimates in standard PRS analysis when non-random sampling is not addressed. In contrast, when selection bias is accounted for using inverse probability (IP) weighting, socioeconomic status (SES) and healthcare utilization are no longer associated with biobank enrollment, and so biobank enrollment is no longer a descendant of a collider. Therefore, stratifying on biobank enrollment would not open the non-causal path blocked by healthcare utilization (represented as a tight faucet in the figure below). Thus, IP-weighted PRS estimates would likely represent effects through the causal path only.

We first evaluated the impact of IP-weighting on the penetrance (i.e., case prevalence as a function of PRS) by comparing the weighted case prevalence against the unweighted case prevalence (34). Next, we evaluated the discrimination of the PRS using the area under the receiver operator characteristic curve (hereafter, the AUC) (35). Under the unweighted approach, we fitted standard logistic regression models adjusting for covariates. Under the IP-weighted approaches, we inputted the standard and modular IP-weights, respectively, and fitted weighted logistic regression models. We then we calculated the AUC to compare performance of the unweighted and IP-weighted logistic regression models (36). Lastly, we estimated the discrimination of psychiatric PRS using the same method in subsamples defined by sex assigned at birth and current age to explore potential effect modification by these factors.

## RESULTS

### Descriptive statistics

As shown in **Table 1**, we first compared participants in the analytic (Biobank) sample (N=24,153) for the PRS analyses against those who were not included (from the broader pool of eligible patients in the healthcare system). In general, the included individuals were significantly more likely to be male, veterans, and married, have publicly funded insurance, and have markedly greater healthcare utilization compared to those excluded and those in the overall source population. Additionally, we compared the prevalence estimates for common health conditions of those included in the final analytic sample against those of excluded participants (see **eTable 1**). Consistent with their higher frequency of healthcare interactions, individuals included in the Biobank PRS analysis were more likely to have clinical diagnoses of all disease conditions examined, including up to three times higher rates of endocrine, nutritional, and metabolic diseases (e.g., Type 1 and 2 diabetes mellitus, obesity), neuropsychiatric conditions (e.g., neurological disorders, major depressive disorder, suicidal behavior), and circulatory conditions (e.g., essential hypertension, myocardial infarction). Of note, the prevalence of rheumatoid arthritis was up to five times greater among those included than those not included in the PRS analyses, likely reflecting recruitment into the MGB Biobank from rheumatology clinics. Lastly, the prevalence estimates for schizophrenia, bipolar disorder, and depression in the analytic sample of 24,153 MGB Biobank participants were 1.0% (n_case_=236), 4.5% (n_case_=1,079), and 26.2% (n_case_=6,329), respectively.

**Table 1.**
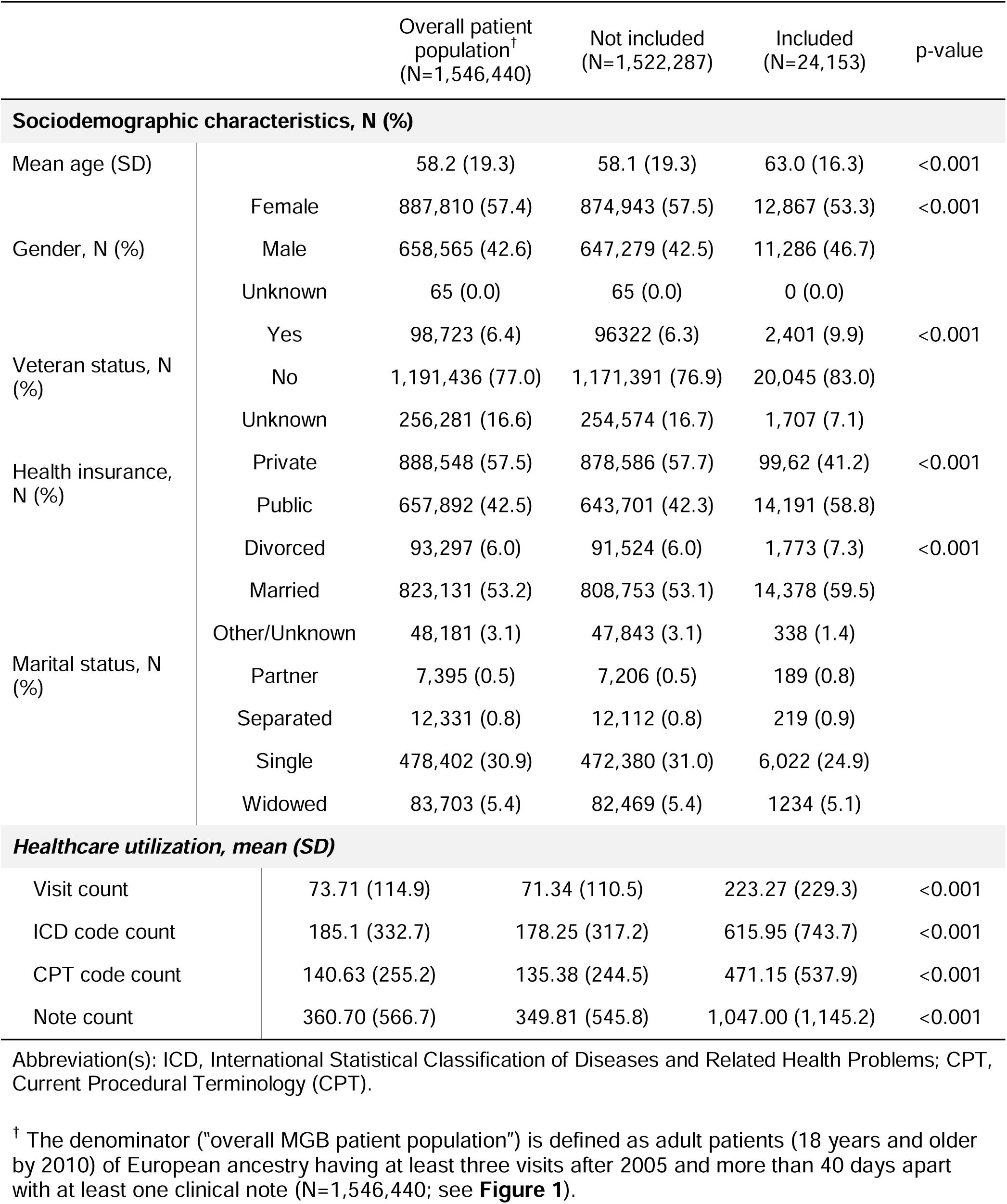
Comparison of demographic and healthcare utilization characteristics of patients self-identifying as non-Hispanic White in the overall MGB patient population^†^ against those included in the PRS analysis (shown in number of participants and prevalence of a given condition).

### Identification of key determinants of selection in the MGB Biobank

In the XGBoost model under the standard IP weighting approach, visit count, note count, current age, and clinical encounters at Massachusetts General Hospital (MGH) or Brigham and Women’s Hospital (BWH) were the five most informative features that differentiated those included and those not included in the PRS analysis, followed by treatment at Northshore Medical Center or Newton-Wellesley Hospital and median neighborhood income in 2010 (see **eFigure 2a**). The top features indicative of healthcare utilization from the standard IP weighting approach also appeared in the three XGBoost models under the modular IP weighting approach. The modular approach identified additional features that contributed to the probability of being retained in each step of selection, such as anxiety, phobic, and dissociative disorders, ischemic heart disease, treatment history at Faulkner Hospital, and rheumatoid arthritis and other inflammatory polyarthropathies (see **eFigures 2b-d**).

In addition to overall feature importance, we further examined the directionality of feature contributions to being retained in each step of selection in the modular IP weighting approach. This was motivated in part by prior work showing that standard IP weighting can lead to biased estimates when a given feature plays a different role in each step of a sequential selection procedure (21,37,38). To address this, we calculated Shapley values at every observed value of each feature across all possible combinations with other features and evaluated whether key features had dynamic contributions across the three selection steps. Interestingly, visit count, which was the most important feature in every step of selection, exhibited different directions of associations with selection probabilities across the three steps (see **eFigure 3b-d**). For example, an increasing number of visits was associated with a *higher* likelihood of providing consent to participate in the Biobank but a *lower* likelihood of being retained in the subsequent steps of selection. This underscores the incomplete information captured by standard IP weighting when there are factors that impact selection probabilities differently across multiple phases of selection.

### Polygenic risk estimation

#### Case prevalence per deciles of standardized residuals of psychiatric PRS

After standardizing PRS by principal components, sex, age, and genotyping microarray, case prevalence for schizophrenia in the top decile of standardized residuals of schizophrenia PRS was 2.7% (2.1-3.3) in the unweighted analysis, and 2.0% (1.2-2.7) in the standard IP weighted analysis (see **Figure 3a**). The unweighted and IP weighted estimates differed more substantially in the case of bipolar disorder: case prevalence of bipolar disorder in the top PRS decile was 10.0% (8.8-11.2) in the unweighted analysis, but only 6.2% (5.0-7.5) when selection bias was accounted for using IP weights (see **Figure 3b**). Finally, case prevalence of depression in the top decile of standardized residuals of depression PRS was 33.5% (31.7-35.4) in the unweighted analysis but was reduced to 28.9% (25.8 – 31.9) after IP weighting (see **Figure 3c**). Results using modular IP weighting based on intermediate selection steps were similar to those observed with standard IP weighting (see **eTables 5-7**).

**Figure 3.**
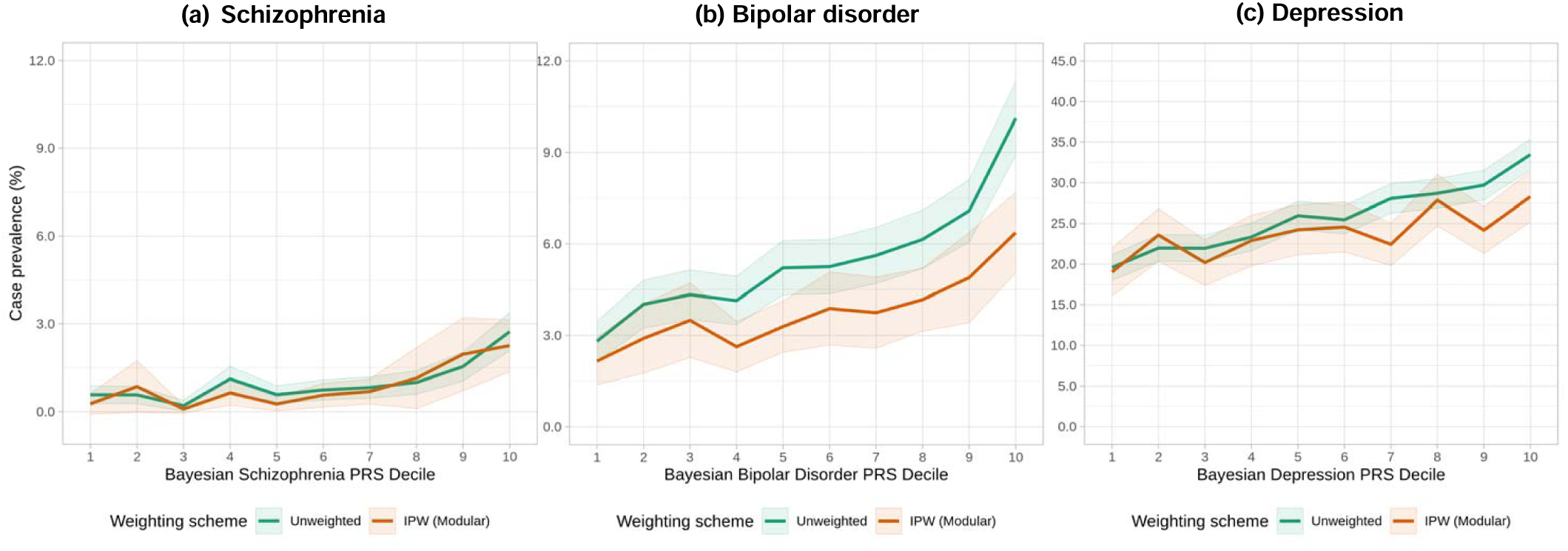
Case prevalence by polygenic risk score (PRS) decile for three psychiatric traits using two different weighting schemes— unweighted and modular IP-weighted. PRS were adjusted for potential confounding by top genetic principal components, sex, age, and genotyping microarray. The solid lines indicate point estimates, and the bands indicate 95% confidence intervals for corresponding point estimates. Note that the standard IP-weighted model is not shown in this figure, since the estimates were nearly identical to the modular IP-weighted model. Numeric estimates from all three models can be found in **eTables 5** through **7**.

#### Discrimination of psychiatric PRS

We found the largest impact of IP-weighting on discrimination with respect to schizophrenia relative to bipolar disorder and depression (see **eTable 8**). When stratified by sex assigned at birth, AUC were generally higher among male participants than female participants regardless of the weighting scheme (see **eFigure 6a)**. The impact of IP-weighting was also greater among males (AUC=0.792 and 0.711 from unweighted and modular IP-weighted models, respectively) than females (AUC=0.711 and 0.675 from unweighted and modular IP-weighted models, respectively).

In addition, we found that both the magnitude and direction of the impact of IP weighting varied by age, especially for schizophrenia (see **eFigure 6b**). For example, among participants whose age was less than 40 years, the AUC of schizophrenia PRS from the unweighted IP-weighted model was lower than the AUC from the modular IP-weighted model. Conversely, the AUC from the unweighted model was higher than the AUC from the modular IP-weighted model among participants whose age was greater than or equal to 40.

## DISCUSSION

As interest grows in returning PRS-based risk estimates to participants in both research and clinical settings, the robustness of such estimates becomes increasingly important. In the present study, we demonstrated that effect estimates of psychiatric PRS can be sensitive to selection bias, using the MGB Biobank as a case example. First, we showed that volunteer-based biobank participants may substantially differ from patients in the underlying healthcare system with respect to a wide range of patient profiles including sociodemographic, healthcare utilization, and clinical characteristics. Notably, prevalence of disease conditions and rates of healthcare utilization were substantially higher in the analytic sample than in the overall MGB patient population. This suggests that, in contrast to the well-known phenomenon of “healthy volunteer bias” (39–43), patients enrolled in hospital-based biobanks may have a *greater* burden of illness than those in the underlying healthcare system from which they were selected. In addition, we demonstrated the utility of an efficient machine learning algorithm to identify demographic and clinical variables that are significantly associated with selection in biobank PRS analyses and to adjust for selection bias.

Using IP weighting procedures, we found that selection bias can produce meaningful effects on estimates of penetrance and discrimination of psychiatric PRS in biobank samples derived from healthcare system populations. Overall, unweighted effect estimates of psychiatric PRS were larger than the IP weighted estimates for the three psychiatric traits examined in the current study. Using the example of a bipolar disorder PRS, **Figure 2** depicts a causal diagram (directed acyclic graph) to illustrate how selection bias might inflate PRS effect estimates in hospital-based biobanks, such as the MGB Biobank. Restriction of PRS analysis to biobank participants is represented as a box around biobank enrollment in the causal diagram. In this example, stratification on the descendent of healthcare utilization, a common effect (i.e., collider) of bipolar disorder PRS and clinical diagnosis of bipolar disorder, can induce a spurious association between the PRS and the target trait—a phenomenon commonly referred to as “collider stratification bias” and known to pose a potential threat to the internal validity (44). As such, the estimated effect could include not only true causal effects but also the spurious association, thereby resulting in larger estimates in standard PRS analysis when non-random sampling is not addressed.

These findings underscore the complex nature of selection bias and the difficulty of predicting the magnitude or direction of the effects by this type of bias on PRS estimates in real-world settings. For example, individuals who are more health-conscious or better informed about the clinical utility of genomic findings may be more willing to participate in a biobank, as has been shown in the UK Biobank (43,45). Conversely, patients whose illness leads to more frequent encounters with the healthcare system may have more opportunities to be selected for biobank participation, leading to an overrepresentation of less healthy individuals. In addition, some individuals may enroll in genetic studies because they have a family history of diseases, such as cancer, and are thus motivated to learn about their risk of illness; enrichment for family history of specific diseases may contribute to differences between biobank cohorts and their underlying source populations.

Recently, several analytic approaches to model and mitigate selection bias in EHR data have been proposed, with varying conceptual definitions of selection bias and statistical approaches to modeling selection mechanisms. For instance, Haneuse and Daniels (20,21) conceptualized selection bias as a missing data problem and encouraged researchers to modularize complex selection mechanisms into a series of sub-mechanisms that are easier to characterize and model. In the current study, we adapted this statistical framework to accommodate the selection procedures unique to PRS analyses conducted in hospital-based biobanks, though results of modular IP weighting did not differ substantially from standard IP weighting in our sample. As an alternative, Goldstein and colleagues (46) proposed controlling for the number of healthcare encounters. However, as they note, stratification on healthcare utilization may actually induce spurious association between two disease phenotypes in cases where healthcare encounters may be the common outcome of the exposure and outcome (i.e., collider stratification bias).

More recently, Beesley and Mukherjee proposed calibration weighting and IP weighting methods to account for selection bias in EHR-linked biobank studies (47). They focus on the form of selection bias that arises from the lack of representativeness and propose constructing weights from external data that better represent the demographic and clinical characteristics of the source population, such as national disease registries for target traits of interest. However, different healthcare systems serve different patient populations, each characterized by unique profiles of sociodemographic, clinical, and healthcare utilization characteristics. As such, it may not be feasible to directly transport selection weight models trained in one healthcare system to another. Instead, adjustment may require a population-specific examination of underlying distributions of the key determinants leading to retention in the analytic sample for PRS analyses. To that end, we leveraged the longitudinal EHRs linked to genomic data collected to derive a set of weights that are specific to the underlying selection mechanism for the MGB Biobank.

Relatedly, different health system biobanks may rely on varying strategies for recruitment and biospecimen collection. For example, at the MGB Biobank, participant enrollment is conducted using a range of procedures including recruitment via a) outpatient primary care or specialty clinics; b) inpatient settings; c) at centralized phlebotomy services; d) online enrollment; or e) collaborating studies. For a subset of patients, biospecimen collection was obtained by placing an order into the EHR (Epic) system to collect a sample concurrently with a clinically ordered blood draw. Although an overrepresentation of less healthy individuals could be a general characteristic of hospital-based biobanks given that they originate from patient populations, the degree of overrepresentation may further vary depending on the distinct method of recruitment and sample collection used in each biobank study.

Our results should be interpreted in light of several limitations. First, our approach does not address another threat to the validity of PRS risk estimates implemented in healthcare settings—the distributional mismatch between the sample in which the PRS is trained and the samples in which the PRS is being validated or implemented. Such estimates are typically derived from validation samples to which allelic weights found in an independent discovery GWAS are applied. To the extent that the discovery and validation samples differ from the implementation sample (here, the MGB biobank) on a range of factors (e.g., age, sex, socioeconomic status) that can affect PRS penetrance, risk estimates may be miscalibrated (48). Second, we examined only three psychiatric traits and our results suggest that the impact of selection bias will vary across different clinical conditions. Lastly, the current investigation was limited to subjects with self-reported white race. Thus, further investigation and validation in other ancestry populations as well as in non-psychiatric conditions are necessary to evaluate the generalizability of our results.

In conclusion, our analyses demonstrate a novel approach for detecting and accounting for unrecognized selection bias in polygenic risk estimation in hospital-based biobank samples. With the growing interest in the return of genomic risk information in clinical practice, it will be important to address such biases to avoid adverse impacts on clinical practice and patient outcomes.

## Data Availability

All data produced in the present study are available upon reasonable request to the authors.

## ACKNOWLEDGEMENTS

This work was conducted with support from Harvard Catalyst | The Harvard Clinical and Translational Science Center (National Center for Advancing Translational Sciences, National Institutes of Health Award UL1 TR002541) and financial contributions from Harvard University and its affiliated academic healthcare centers. The content is solely the responsibility of the authors and does not necessarily represent the official views of Harvard Catalyst, Harvard University and its affiliated academic healthcare centers, or the National Institutes of Health.

YAF is supported by the National Taiwan University Higher Education Sprout Project (NTU-110L8810) within the framework of the Higher Education Sprout Project by the Ministry of Education (MOE) in Taiwan. EWK was supported by 5U01HG008685. TG was supported in part by NIA R00AG054573, NHGRI U01HG008685 and NHGRI U01HG011723. JWS was supported in part by NIMH R01MH118233, NHGRI U01HG008685, and a gift from the Demarest Lloyd, Jr. Foundation.

This study would not be possible without the contributions of Mass General Brigham (MGB) patients and Biobank participants. We would also like to thank the research coordinators and the Biobank study for their tremendous effort in participant recruitment and sample collection. Lastly, we would like to acknowledge the RPDR team for their work maintaining the enterprise research patient data warehouse.

## DISCLOSURES

Dr. Smoller is a member of the Leon Levy Foundation Neuroscience Advisory Board, the Scientific Advisory Board of Sensorium Therapeutics, and has received honoraria for internal seminars at Biogen, Inc and Tempus Labs. He is PI of a collaborative study of the genetics of depression and bipolar disorder sponsored by 23andMe for which 23andMe provides analysis time as in-kind support but no payments.

## Notes

### Author Declarations

This research was conducted as part of the PsycheMERGE Consortium, under approval from the Mass General Brigham Institutional Review Board.

