## Supplementary material for "Impact of Selection Bias on Polygenic Risk Score Estimates in Healthcare Settings": Online Supplemental Methods and Figures

**eMethod.** Supplemental methods.

**eFigure 1.** Visualization of the two different inverse probability (IP) weighting approaches accounting for two different selection schemes: (a) standard and (b) modular approaches.

**eFigure 2.** Feature importance of top 20 features from the standard and modular IP weight models using the mean absolute Shapley values.

**eFigure 3.** SHAP beeswarm plots showing directionality and magnitude of effects on the selection probabilities by the top 20 features in the corresponding IP weight models.

**eFigure 4.** Case prevalence per deciles of standardized residuals of PRS, stratified by sex assigned at birth.

**eFigure 5.** Case prevalence per deciles of standardized residuals of PRS, stratified by current age.

**eFigure 6.** Comparison of discrimination by psychiatric PRS across groups defined by sex assigned at birth and current age.

**eTable 1-8 can be found in the accompanying Excel spreadsheets.**

This supplementary material has been provided by the authors to give readers additional information about their work.

**eMethods**

**Genomic data processing**

The MGB Biobank samples were genotyped on Multi-Ethnic Global array (MEGA) from Illumina (Illumina Inc., San Diego, USA) and released in several batches. We performed batch-specific genotype data QC to remove single nucleotide polymorphisms (SNPs) with genotype missing rate >0.05, samples with genotype missing rate >0.02, and SNPs with differential missing rate >0.01 between any two batches, after which different batches were merged for subsequent QC steps. As MGB Biobank included individuals from diverse populations, we inferred genetic ancestry of biobank participants using 1000 Genomes samples (1KG) as the population reference panel (1). Specifically, we computed principal components (PCs) for biobank samples and 1KG samples combined and trained a random forest classifier to assign a “super population” label for biobank samples with a prediction probability ≥0.9 using the first 6 PCs of the 1KG samples as the training data. This resulted in 26,677 individuals whose ancestry was classified as European (EUR), 1,607 as African (AFR), 1,840 as Admixed American (AMR), 504 as East Asian (EAS) and 297 as South Asian (SAS) ancestry. Within each ancestry, we removed samples with a mismatched reported and genetic sex, outliers of the absolute value of heterozygosity (>5 standard deviations from the mean), and one from each pair of related individuals (identity-by-descent (IBD) >0.2); SNPs that showed significant batch associations at P < 1 × 10−4, had a missing rate > 0.02 or Hardy–Weinberg equilibrium (HWE) test P < 1 × 10−10 were also discarded. Next, we used the Michigan Imputation Server (Minimac4) to impute genotype dosages for biobank samples, with the Haplotype Reference Consortium (HRC) as the reference panel for EUR ancestry. Lastly, we removed markers with imputation quality INFO score <0.8, minor allele frequency (MAF) <0.01, a significant deviation from HWE with P < 1 × 10−10, and missing rate >0.02. The dataset uses genome build 37 (hg19). Further information about genotyping, QC, imputation, and population assignment procedures for the MGB Biobank is available on the GitHub repository (<https://github.com/Annefeng/PBK-QC-pipeline>).

**Construction of Bayesian polygenic risk scores (PRS)**

We generated PRS for the European ancestry participants of the MGB Biobank using PRS-CS-Auto (2), a Bayesian polygenic prediction method, based on their genotype data and publicly available summary statistics from the largest, European ancestry genome-wide association studies (GWAS) of schizophrenia (3), bipolar disorder (4), and depression (5). PRS-CS-Auto places a continuous shrinkage (CS) prior on SNP effect sizes and infers posterior SNP weights using GWAS summary statistics and an external linkage disequilibrium (LD) reference panel (e.g., 1000 Genomes Project European Samples). Allowing multivariate modeling of local LD patterns, PRS-CS-Auto is robust to diverse underlying genetic architectures and can increase the accuracy of PRS over conventional approaches (2). We generated PRS for each individual by summing all risk-associated variants weighted by their posterior effect size estimates inferred by PRS-CS-Auto using PLINK, version 2.0 (6). In addition to the continuous PRS, we calculated deciles of the standardized residuals of PRS after adjusting for top 20 ancestry PCs, sex, age, and genotyping microarray.

**eFigure 1.** Visualization of the two different inverse probability (IP) weighting approaches accounting for two different selection schemes: (a) standard and (b) modular approaches.

(a) **Standard IP weighting approach**

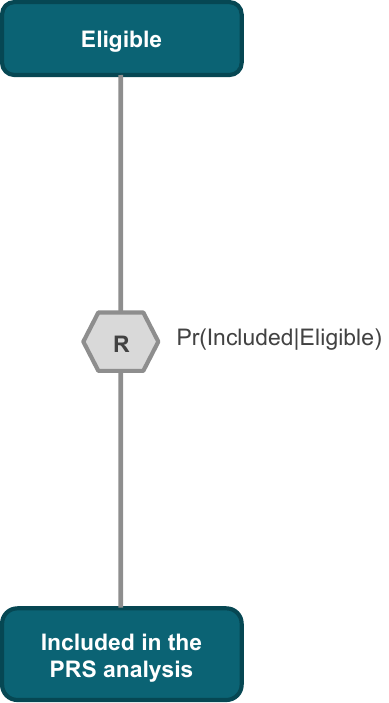

(b) **Modular IP weighting approach**

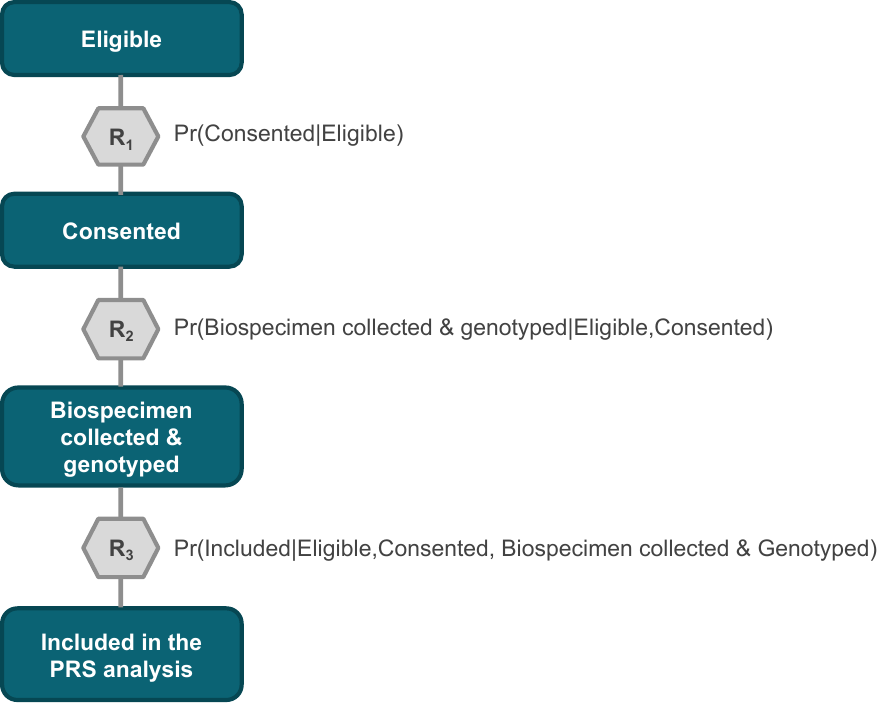

**eFigure 2.** Evaluation of the marginal contribution by top 20 features from the standard and modular IP weight models based on Shapley values. Features having higher mean absolute Shapley values would have a more significant impact on the model’s decision than those having lower values. On the vertical axis, features are rank-sorted based on the magnitude of the mean absolute Shapley values from high (top) to low (bottom).

(a) **Standard IPW**: Pr(Included|Eligible)
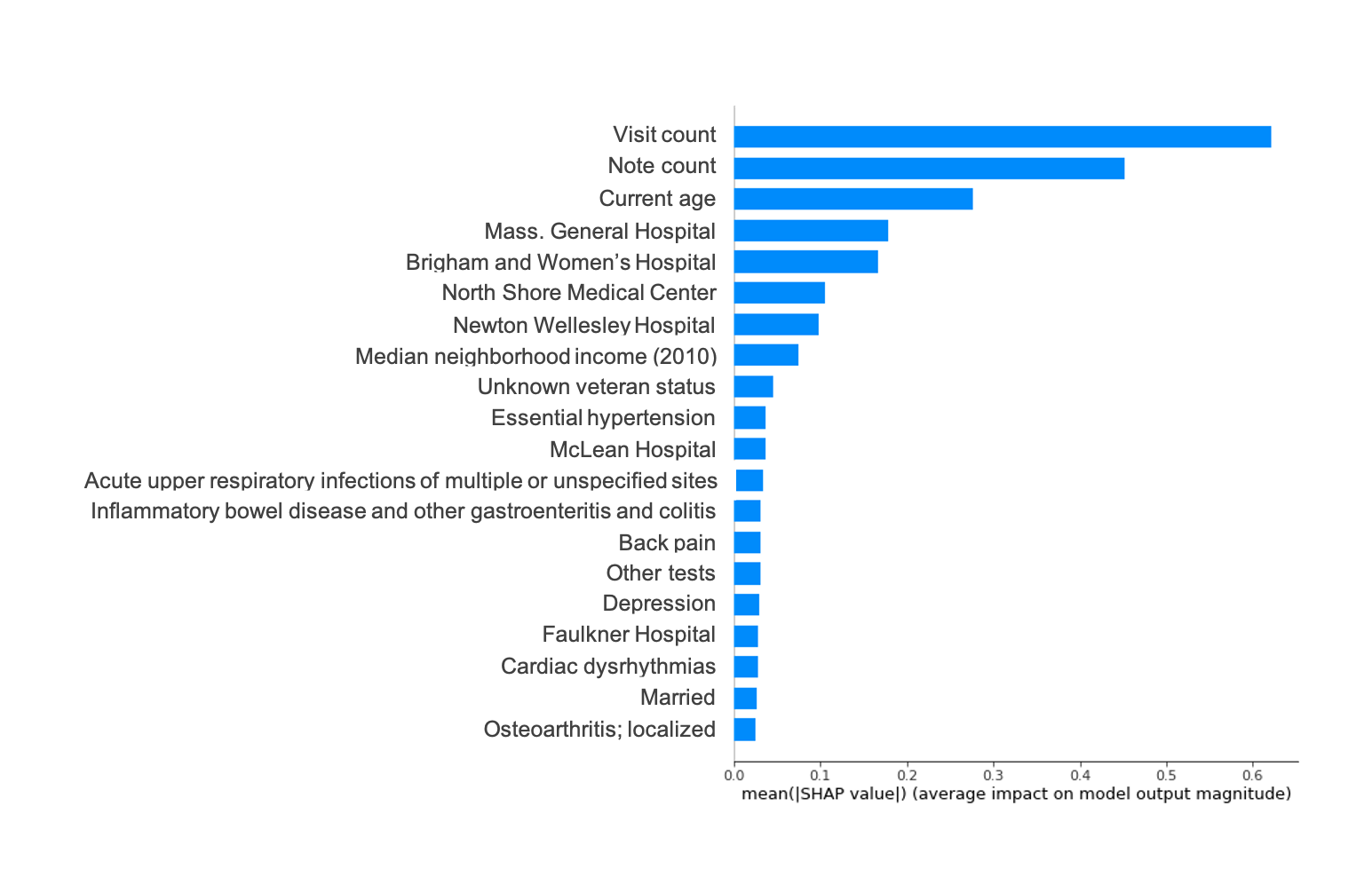

(b) **Modular IPW** – Step 1: Pr(Consented|Eligible)
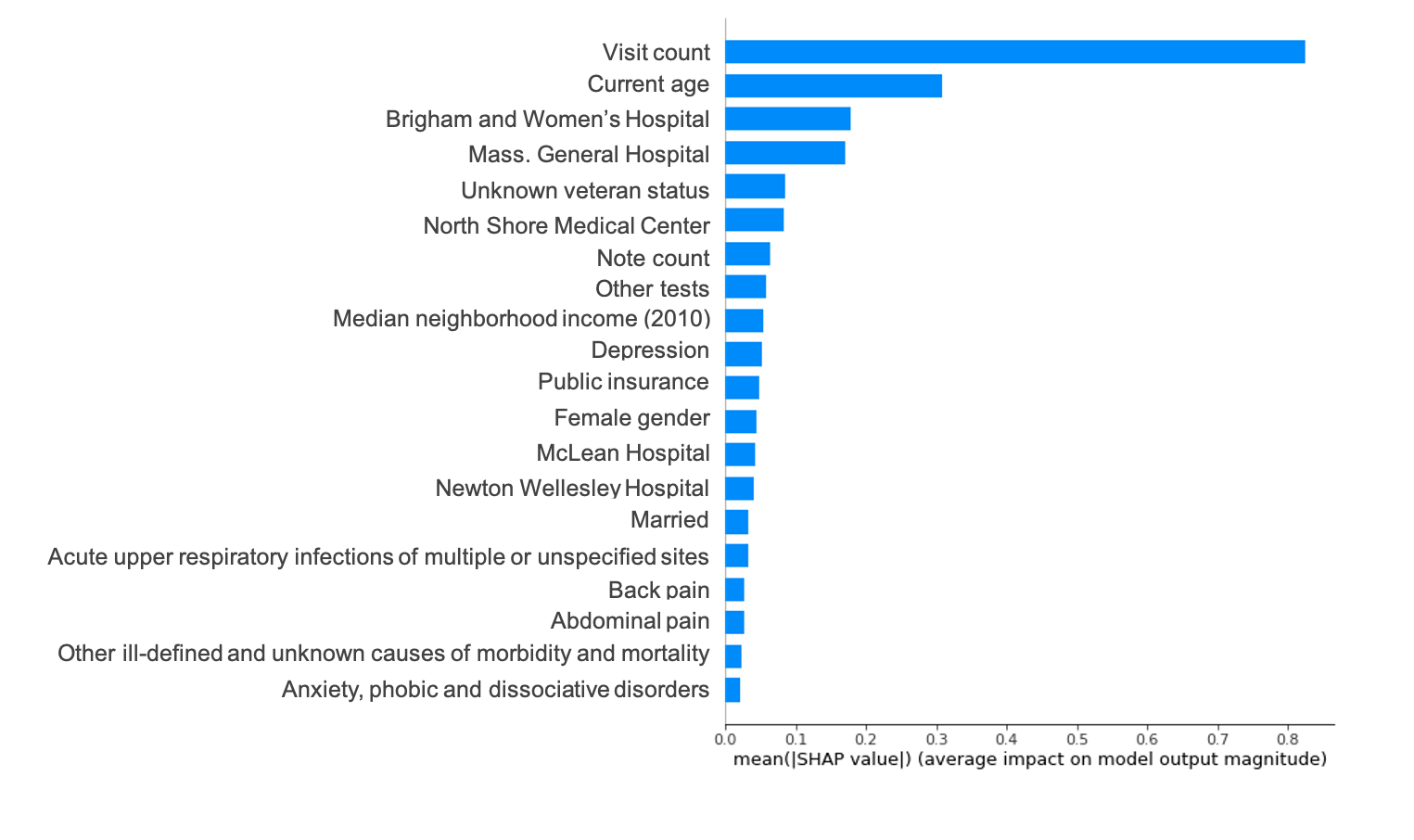

(c) **Modular IPW** – Step 2: Pr(Biospecimen genotyped|Eligible,Consented)
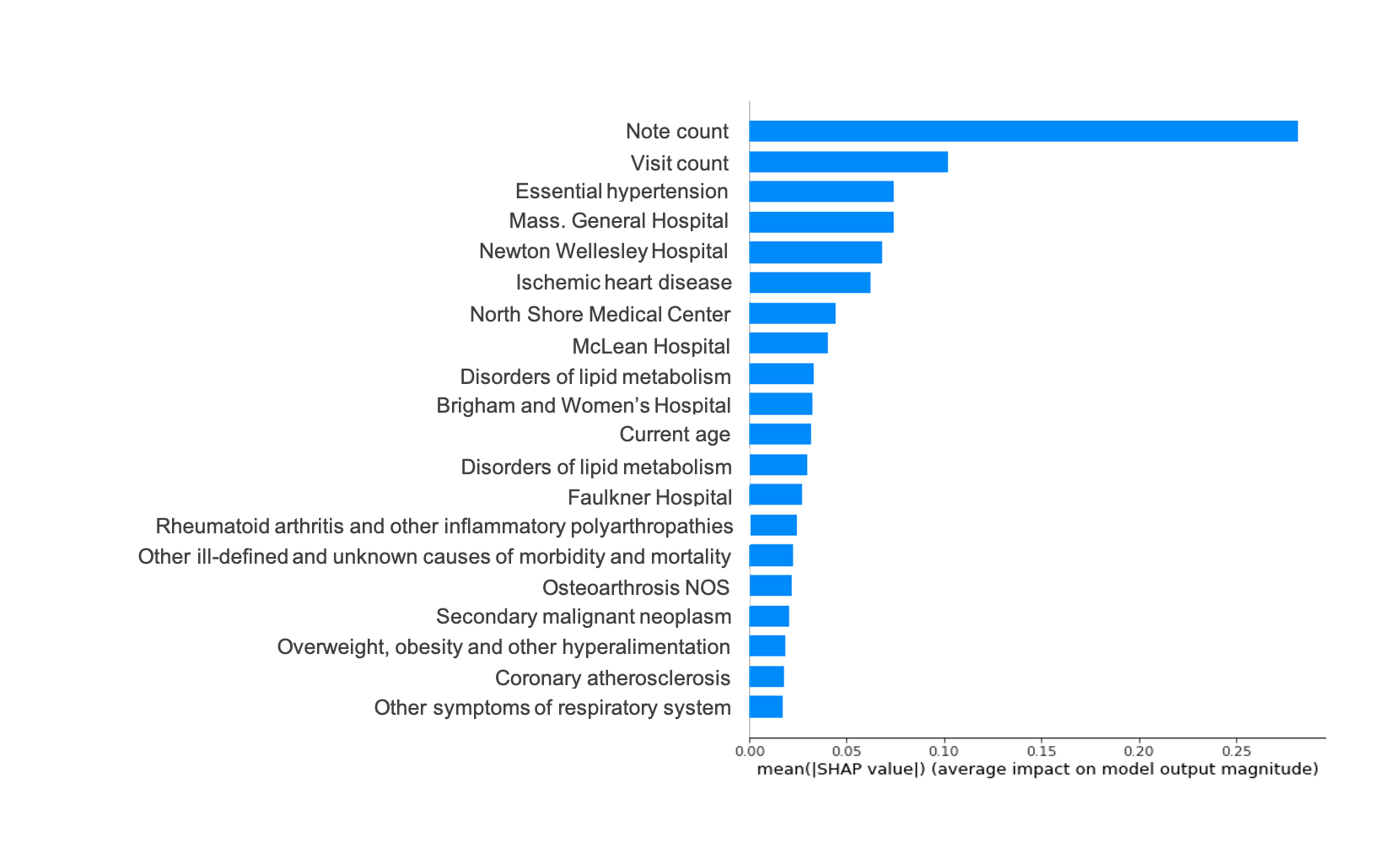

(d) **Modular IPW** – Step 3: Pr(Included|Eligible,Consented,Biospecimen genotyped)
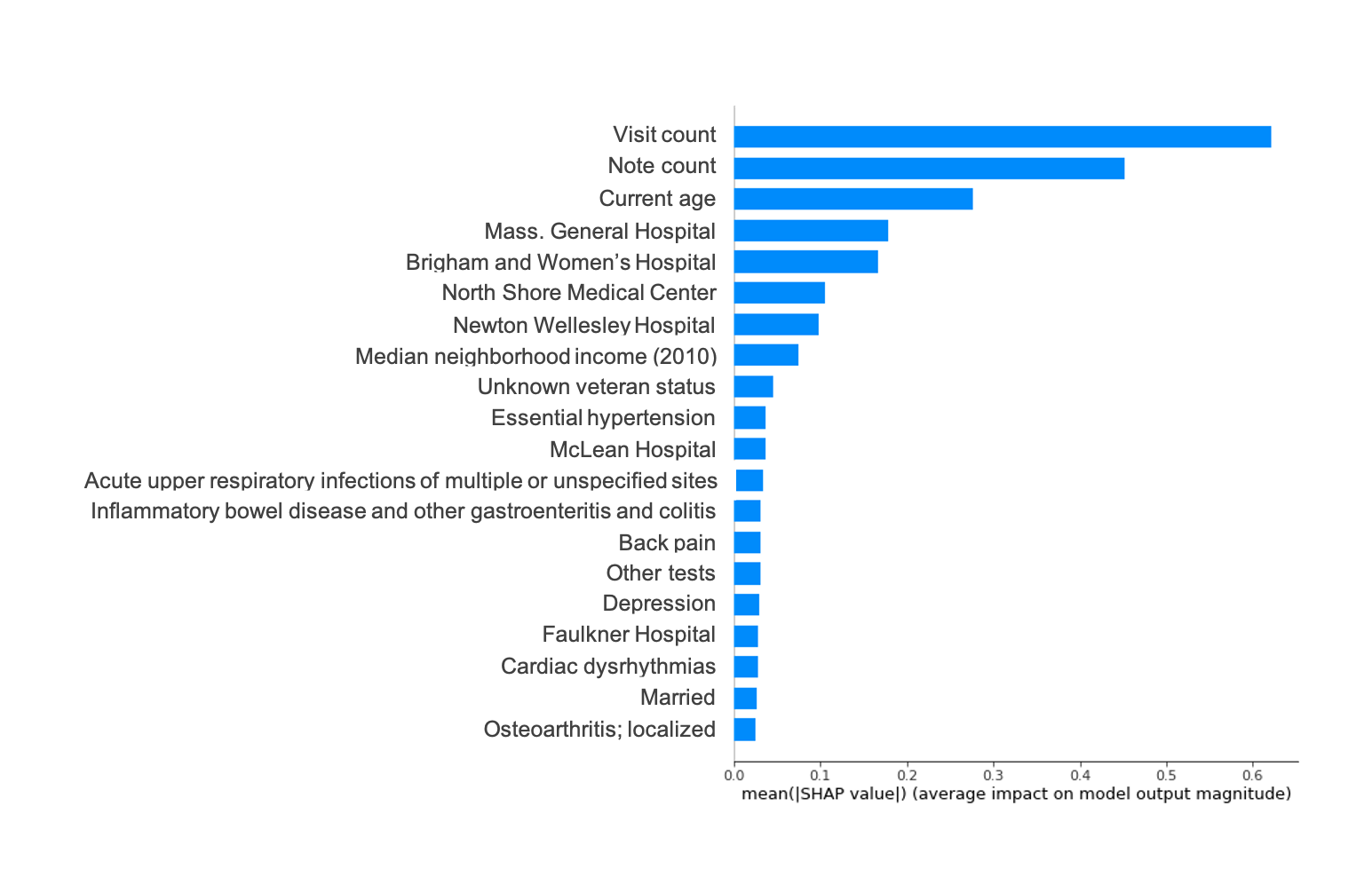

**eFigure 3**. SHAP beeswarm plot showing directionality of effects for the top 20 features in the corresponding IP weight models. The features are rank-sorted in the same order as in the eFigure 2 with the `visit count` feature having the highest mean absolute Shapley values (i.e., most important feature in the standard IP-weighted model). The position of dots on the horizontal axis is determined by the Shapley value of that feature (averaged across all participants), and dots pile up along each feature row to show density. The color of dots is used to display the original value of a feature (i.e., higher feature values would be redder).

(a)
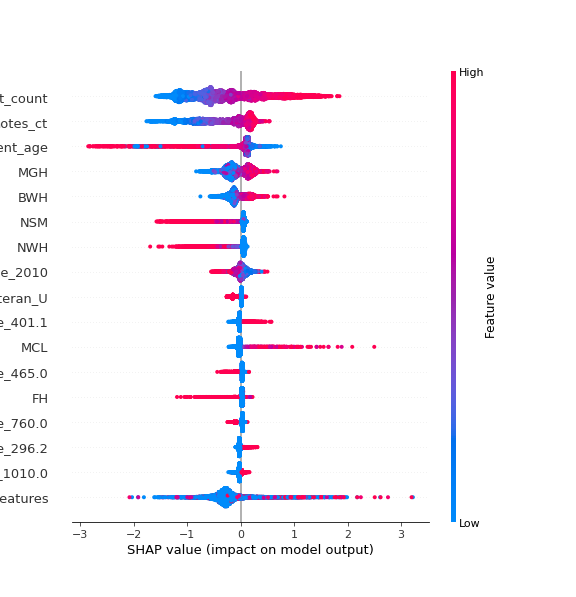
**Standard IP weighting approach** – Pr(Included|Eligible)

Visit count

Notes count

Current age

Massachusetts General Hospital

Brigham & Women’s Hospital

North Shore Medical Center

Newton-Wellesley Hospital

Median neighborhood income (2010)

Unknown veteran status

Essential hypertension

McLean Hospital

Acute upper respiratory infection

Faulkner Hospital

Back pain

Depression

Other tests

Other features combined

(b)
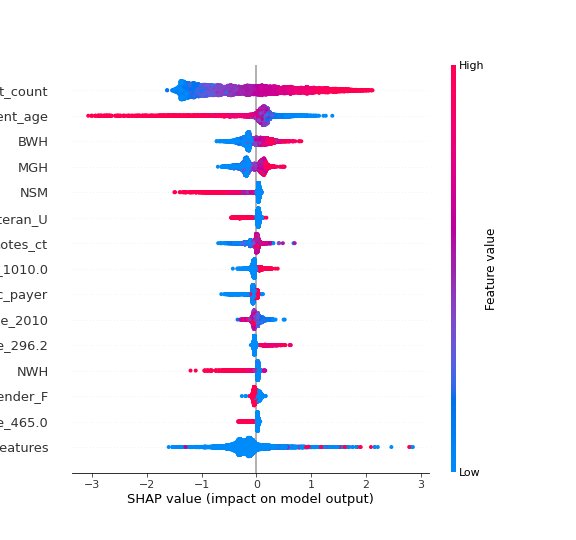
**Modular IP weighting approach** – Step 1: Pr(Consented|Eligible)

Visit count

Current age

Brigham & Women’s Hospital

Massachusetts General Hospital

North Shore Medical Center

Unknown veteran status

Notes count

Other tests

Public insurance

Median neighborhood income (2010)

Depression

Newton-Wellesley Hospital

Female gender

Acute upper respiratory infection

Other features combined

(c)
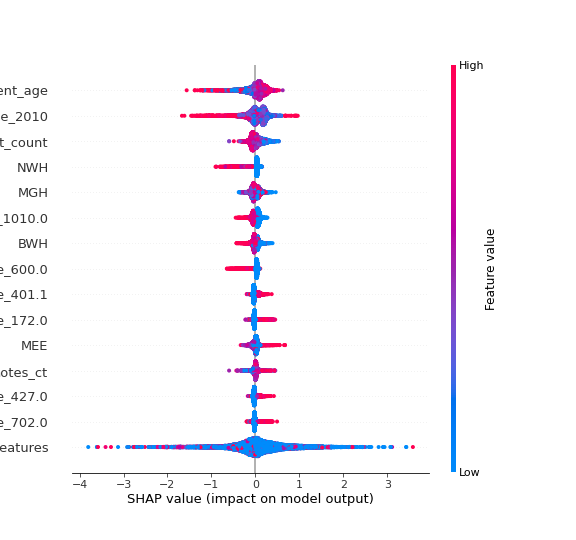
**Modular IP weighting approach** – Step 2: Pr(Biospecimen genotyped| Eligible,Consented)

Current age

Median neighborhood income (2010)

Visit count

Newton-Wellesley Hospital

Massachusetts General Hospital

Other tests

Brigham & Women’s Hospital

Hyperplasia of prostate

Essential hypertension

Skin cancer

Mass Eye and Ear

Notes count

Cardiac dysrhythmias

Degenerative skin conditions and other dermatoses

Other features combined

(d)
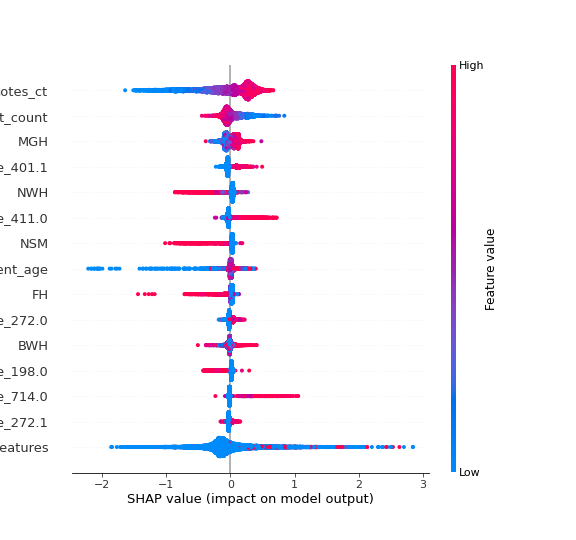
**Modular IP weighting approach** – Step 3: Pr(Included|Eligible,Consented, Biospecimen genotyped)

Notes count

Visit count

Massachusetts General Hospital

Essential hypertension

Newton-Wellesley Hospital

Ischemic heart disease

North Shore Medical Center

Current age

Faulkner Hospital

Disorders of lipoid metabolism

Brigham & Women’s Hospital

Secondary malignant neoplasm

Rheumatoid arthritis and other inflammatory polyarthropathies

Hyperlipidemia

Other features combined

**eFigure 4.** Case prevalence by polygenic risk scores (PRS) decile for three psychiatric traits using two different weighting schemes, stratified by sex assigned at birth. PRS were adjusted for potential confounding by population stratification, sex, age, and genotyping microarray (see **eTable 7a-c** for numeric estimates used to generate the respective figure). The solid lines indicate point estimates, and the bands indicate 95% confidence intervals for corresponding point estimates.

(a) **Schizophrenia**

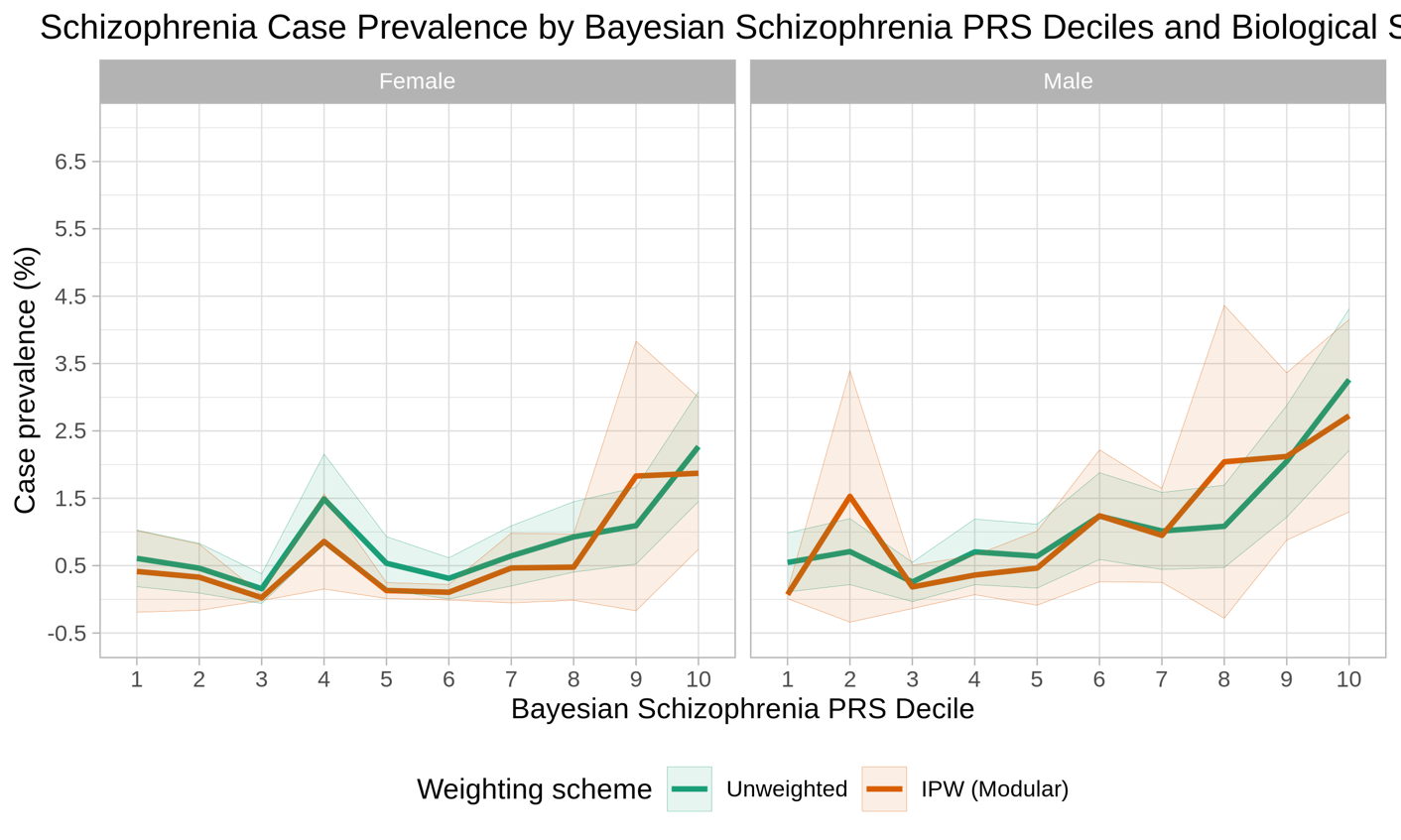

(b) **Bipolar disorder**

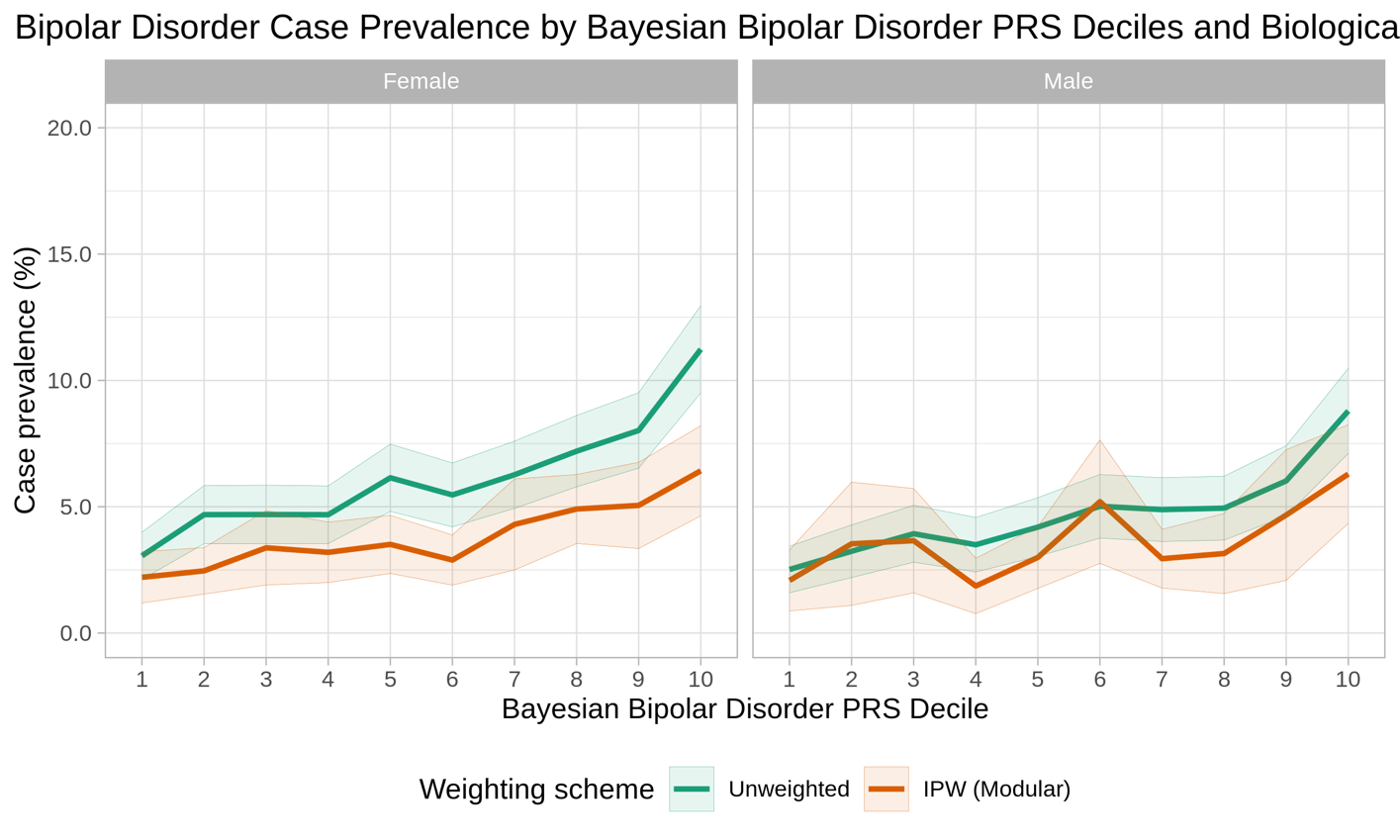

(c) **Depression**

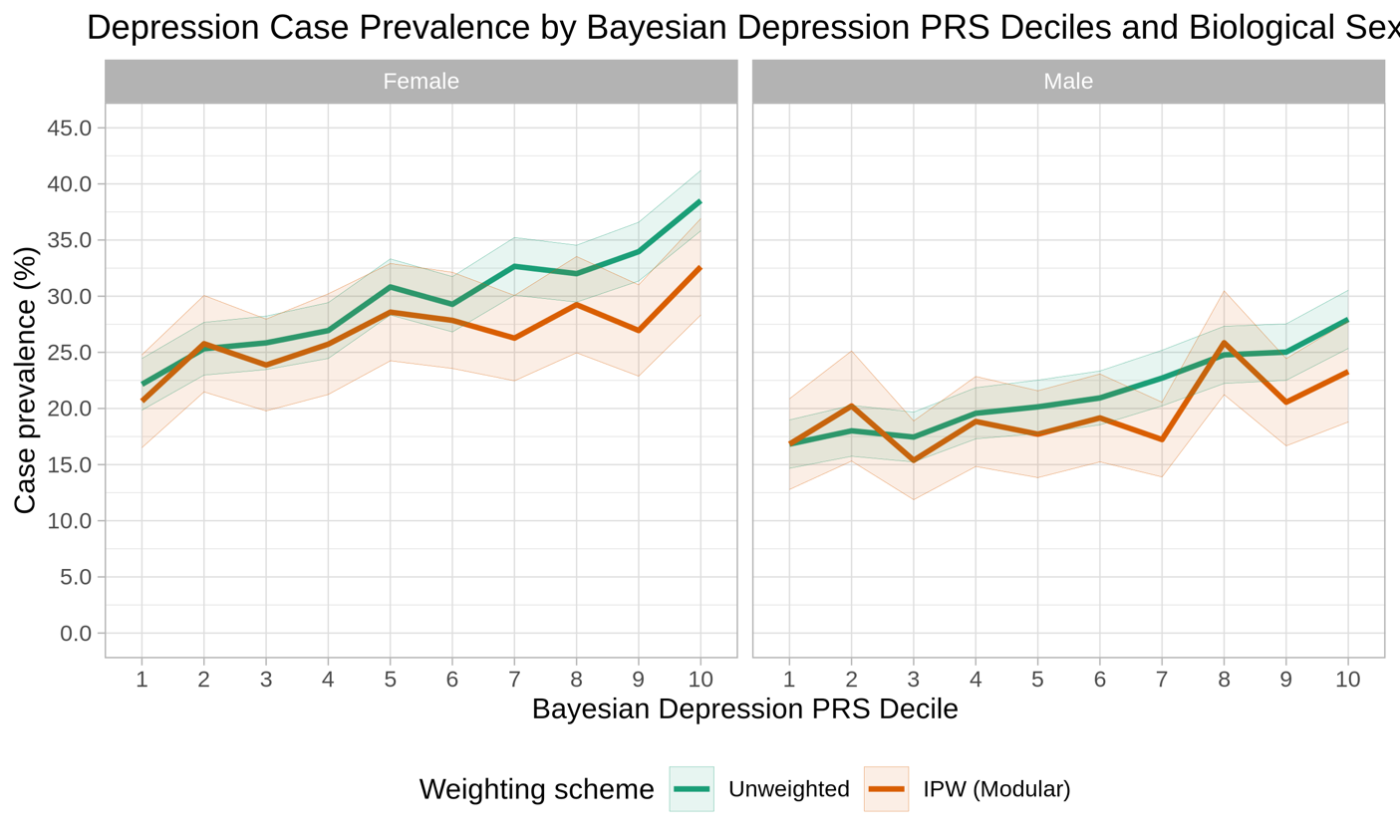

**eFigure 5.** Case prevalence by polygenic risk scores (PRS) decile for three psychiatric traits using two different weighting schemes, stratified by current age. PRS were adjusted for potential confounding by population stratification, sex, age, and genotyping microarray (see **eTable 7a-c** for numeric estimates used to generate the respective figure). The solid lines indicate point estimates, and the bands indicate 95% confidence intervals for corresponding point estimates.

(a) **Schizophrenia**

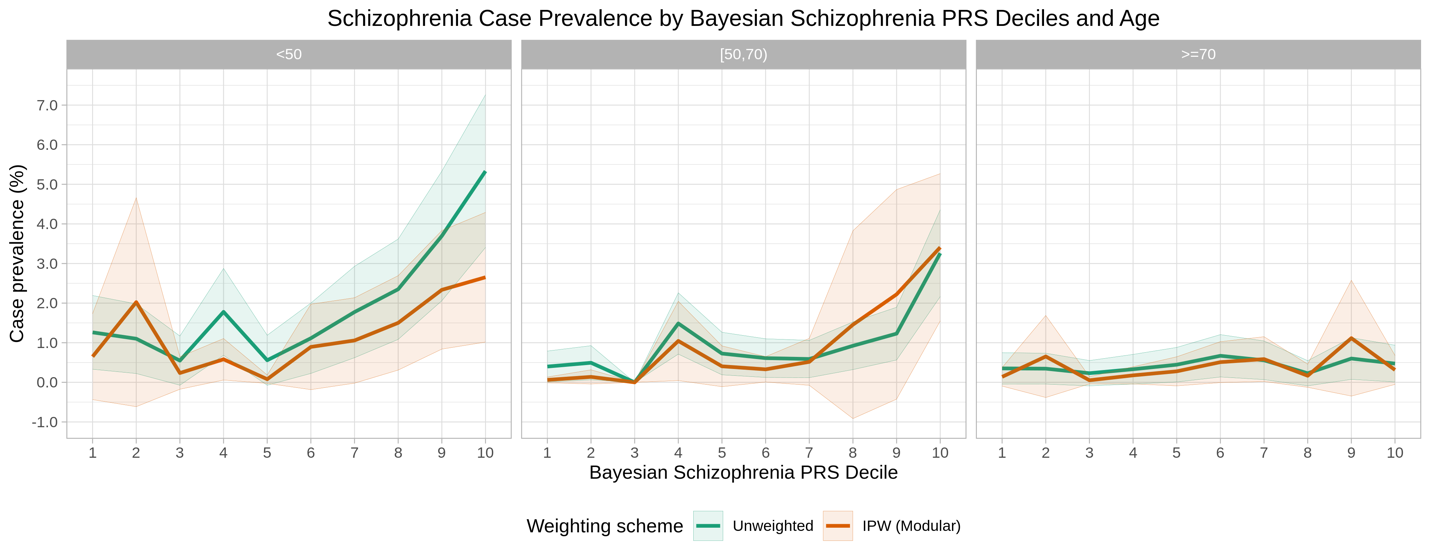

(b) **Bipolar disorder**

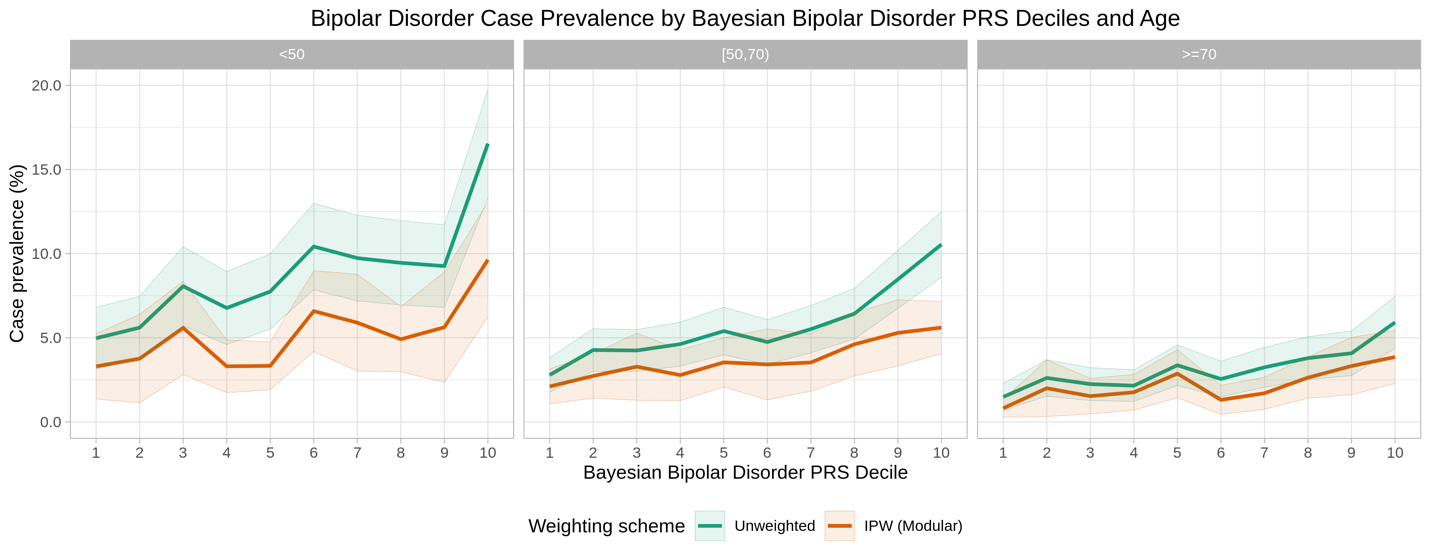

(c) **Depression**

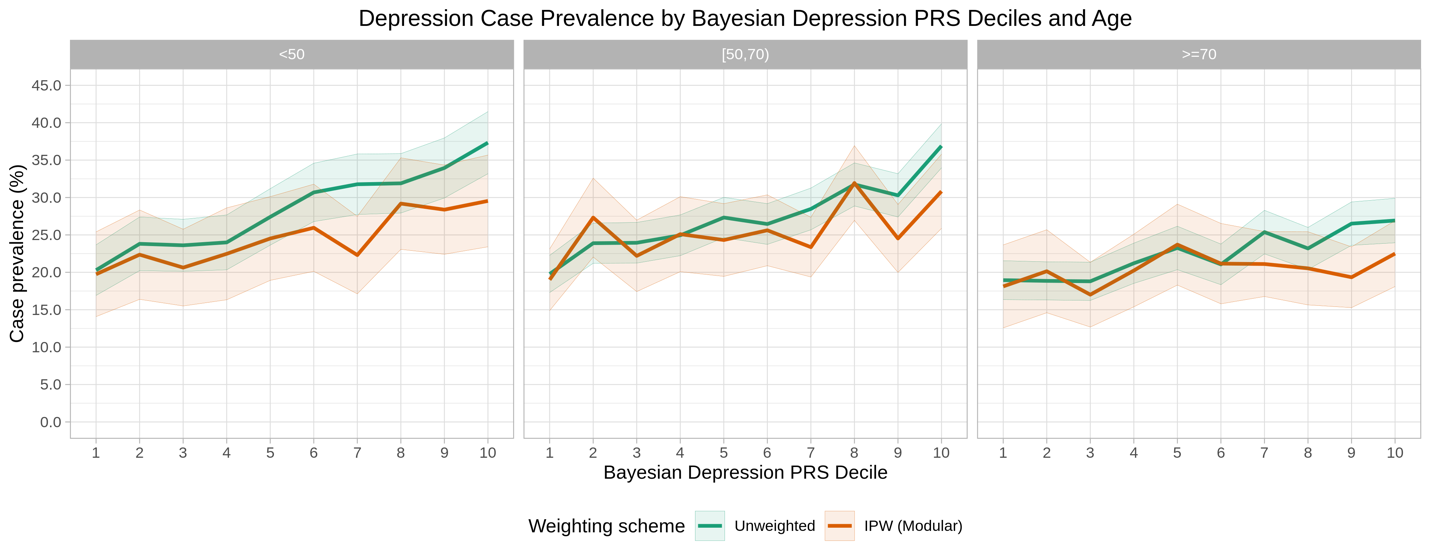

**eFigure 6.** Comparison of discrimination by psychiatric PRS (area under the receiver operating characteristic curve or AUC) across groups defined by sex assigned at birth and age.

(a) **Stratified by sex**

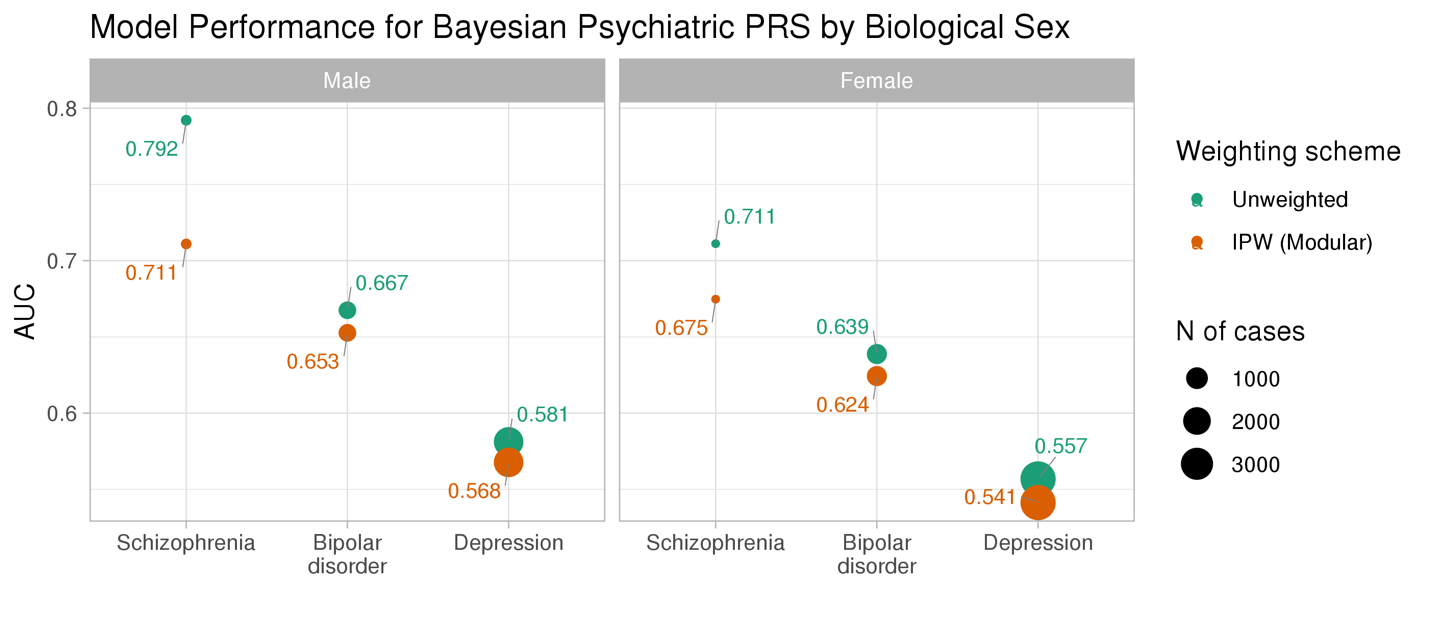

(b) **Stratified by age**

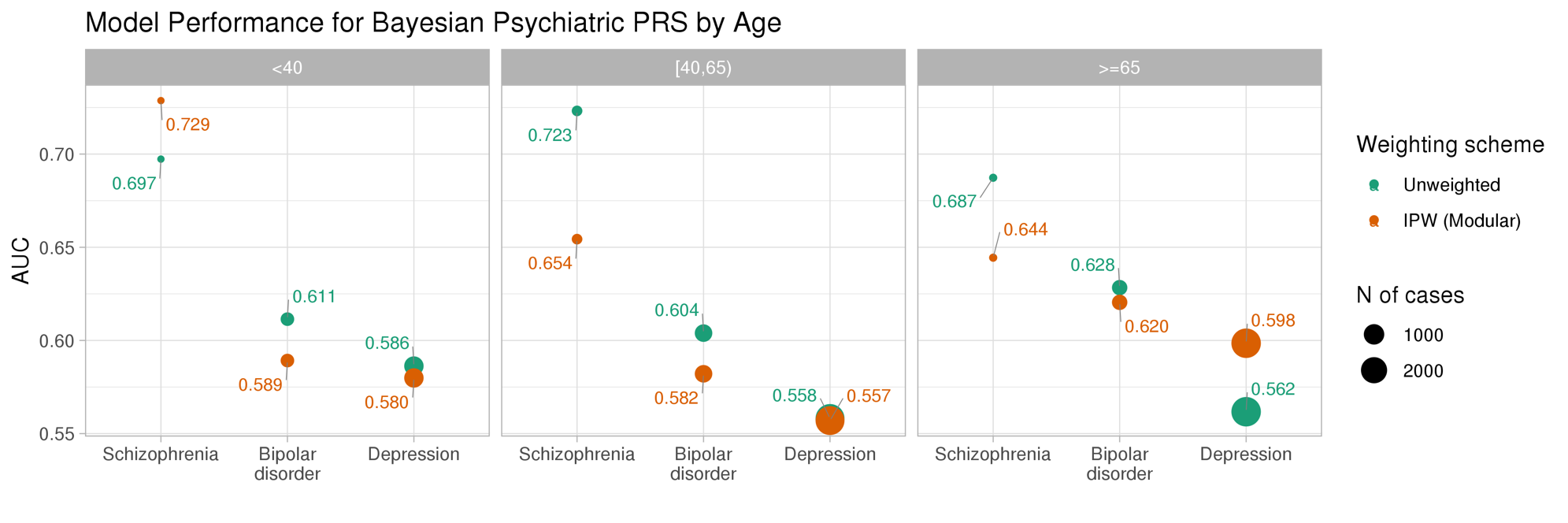
